# Maternal and offspring genome-wide association study of C-reactive protein reveals limited polygenic association with gestational diabetes mellitus

**DOI:** 10.1101/2025.09.30.25336984

**Authors:** Yu Zhang, Amy Moore, Kelli K. Ryckman, Qi Yan, Rafael F. Guerrero, Ming Li, Robert M. Silver, Lynn M Yee, Uma M. Reddy, Maisa N. Feghali, Judith Chung, David M. Haas, Kok Lim Kua, Nianjun Liu

**Affiliations:** Department of Epidemiology and Biostatistics, Indiana University School of Public Health-Bloomington, Bloomington, Indiana, United States of America; Division of Biostatistics and Epidemiology, RTI International, Atlanta, Georgia, United States of America; Department of Obstetrics and Gynecology, Columbia University, New York, New York, United States of America; Department of Biological Sciences, North Carolina State University, Raleigh, North Caroline, United States of America; Division of Maternal-Fetal Medicine, Department of Obstetrics and Gynecology, University of Utah Health Sciences Center, Salt Lake City, Utah, United States of America; Division of Maternal-Fetal Medicine, Department of Obstetrics and Gynecology, Northwestern University Feinberg School of Medicine, Chicago, Illinois, United States of America; Department of Obstetrics, Gynecology and Reproductive Sciences, University of Pittsburgh School of Medicine, Pittsburgh, Pennsylvania, United States of America; Division of Maternal-Fetal Medicine, Department of Obstetrics and Gynecology, University of California, Irvine, School of Medicine, Irvine, California, United States of America; Department of Obstetrics and Gynecology, Indiana University School of Medicine, Indianapolis, Indiana, United States of America; Department of Pediatrics, Division of Neonatal-Perinatal Medicine, Indiana University School of Medicine, Indianapolis, Indiana, United States of America

**Keywords:** early pregnancy, maternal, offspring, CRP, GWAS, PRS, GDM

## Abstract

**Background:** C-reactive protein (CRP) is a well-established biomarker of systemic inflammation. In pregnancy, several studies show association of elevated CRP with gestational diabetes mellitus (GDM). However, the genetic contributions of CRP levels during early pregnancy and their potential association with GDM remain largely understudied.

**Methods:** We conducted maternal and offspring genome-wide association studies (GWAS) of first-trimester CRP levels in the Nulliparous Pregnancy Outcomes Study: Monitoring Mothers-to-Be (nuMoM2b) cohort. Genetic correlation between CRP and GDM was assessed using linkage disequilibrium score regression (LDSC), and polygenic risk scores (PRS) of CRP were further evaluated for associations with GDM.

**Results:** In the European maternal sub-cohort of nuMoM2b, three genome-wide significant loci (*CRP*, *LEPR*, and *HNF1A*) were associated with early pregnancy CRP, consistent with previous GWAS. Multi-ancestry maternal GWAS revealed two additional associated loci (*ENSG00000257703* and *APOC1*). Offspring GWAS did not identify any genome-wide significant associations. LDSC using the nuMoM2b European maternal CRP GWAS showed no significant genetic correlation with GDM. In contrast, a significant correlation was observed using the large population-based CRP GWAS, suggesting context-specific genetic architecture. However, PRS of CRP, based on GWAS summary statistics from either study, was not significantly associated with GDM risk in the nuMoM2b cohort.

**Conclusions:** Our findings validate known genetic loci regulating systematic inflammation in a cohort of pregnant individuals, correlating with serum CRP levels, and highlight the value of pregnancy-specific GWAS in uncovering unique biological pathways relevant to maternal-fetal health.

**Author summary:** Inflammation plays an important role in pregnancy and has been linked to conditions like gestational diabetes. C-reactive protein (CRP) is a widely used marker of inflammation, but how the genetic makeup of pregnant individuals and their offspring influence maternal CRP levels during early pregnancy is not well understood. We studied the genetic factors affecting CRP levels in over 4,000 pregnant women and 2,000 babies from a large, multi-ancestry U.S. pregnancy study. We found that genes linked to inflammation in the general population, such as CRP, LEPR, HNF1A, APOE, and APOC1 previously, also influence maternal CRP levels during early pregnancy. While CRP levels have been linked to gestational diabetes, we found no strong genetic connection between CRP and GDM. Our study provides important new insight into how genes influence inflammation during pregnancy and offers a foundation for future research in maternal and child health.

## Introduction

C-reactive protein (CRP) is a widely recognized biomarker of systemic inflammation, extensively studied for its role in predicting cardiovascular diseases, metabolic disorders, and adverse pregnancy outcomes [1–5]. During pregnancy, maternal immune and inflammation processes are tightly regulated to support fetal development [6,7]. Disruption of this balance, especially during early pregnancy, can elevate CRP levels, which have been associated with an increased risk of gestational diabetes mellitus (GDM) [8,9], preeclampsia [10], and preterm birth [11–13]. These conditions pose significant long-term health risks to both mother and child, including higher likelihood of cardiovascular and metabolic diseases [11,14]. Understanding how genetic variants influence CRP levels during pregnancy may offer potential avenues for clinical intervention and improving maternal-fetal health.

Several genome-wide association studies (GWAS) have identified genetic variants associated with CRP levels in general populations [15–18]. However, the genetic variants influencing CRP levels during early pregnancy remain largely understudied. This gap is critical, as early pregnancy represents a unique physiological and immunological state that is different from non-pregnant individuals and therefore may require unique genetic regulation of inflammation and CRP production. Prior research showed strong phenotypic associations between higher CRP levels during early pregnancy and increased risk of GDM [19]. However, it remains unclear whether genetic variants that influence CRP levels also contribute to GDM risk.

In addition to maternal genetic influences, fetal genetic variations may also contribute to maternal physiological traits during pregnancy. This phenomenon, sometimes referred to as “fetal drive,” hypothesizes that genetic variants inherited from the fetus can influence maternal outcomes such as blood pressure and metabolic regulation [20–23]. Prior studies have demonstrated associations between fetal genetic variations and maternal blood pressure after adjusting for maternal genotype, supporting the idea that fetal genes can exert systemic effects on the maternal environment. Whether fetal genetic variation similarly affects maternal CRP during early pregnancy, however, has not yet been investigated.

To address these gaps, we investigated the genetic contributions to CRP levels measured during early pregnancy using maternal genome-wide genotype data from the Nulliparous Pregnancy Outcomes Study: Monitoring Mothers-to-Be (nuMoM2b) cohort [24]—a large, multi-center study of nulliparous women with comprehensive clinical, biomarker, and genomic data. Previous research within the nuMoM2b cohort demonstrated a strong phenotypic association between elevated early pregnancy CRP levels and subsequent risk of GDM [11,25]. To evaluate the potential genetic link between CRP and GDM, we performed linkage disequilibrium score regression (LDSC) to assess shared genome-wide genetic architecture and constructed polygenic risk scores (PRS) for CRP to test whether maternal genetic predisposition to higher CRP levels was associated with increased risk of GDM. Finally, given the potential influence of offspring genetic variation on maternal physiology [23,25], we conducted an additional GWAS of maternal CRP levels using offspring cord blood-derived genotypes, to explore whether offspring genetic variations contribute to maternal inflammation during early pregnancy.

## Materials and methods

### Study population

The study was conducted within the nuMoM2b cohort, a large, multicenter, prospective study that enrolled 10,038 nulliparous pregnant women between 2010 and 2013 at eight clinical sites across the United States [24]. Participants were enrolled during the first trimester of pregnancy (gestational age 6 to13+6 weeks), and comprehensive data collection included demographic information, clinical assessments, and biospecimen sampling. A total of 4,502 maternal serum samples was collected during this visit and processed under standardized protocols in a centralized biorepository to ensure sample integrity. Serum CRP concentrations during the first trimester (mg/L) were measured using the Beckman Coulter AU System High-Sensitivity C-Reactive Protein assay (Reagent OSR6199) on the AU480 Chemistry Analyzer (Beckman Coulter, Brea, CA, USA). Hereafter, we refer to these measurements as first-trimester CRP. As the CRP levels exhibited a left-skewed distribution, a log transformation was applied to normalize the values for subsequent analysis.

Genome-wide genotyping was conducted on 9,757 mothers with whole blood sample using the Infinium Multi-Ethnic Global D2 BeadChip (Illumina, Miami, FL, USA) [26]. Quality control (QC) and genotype imputation were applied, following the pipeline detailed in Yan et al. (2023) [27]. QC procedures included checking for genotyping call rates, Hardy-Weinberg equilibrium, and minor allele frequency. A total of 9,742 women had genotying data that passed QC [27], and 6,669,023 common variants (minor allele frequency (MAF) > 0.05) were available for analysis. The final dataset used for analysis included 4,326 mothers with both genetic data and CRP measurements available. Genome-wide genotyping was performed on 3,860 offspring using DNA extracted from cord blood collected at birth, with the Illumina Multi-Ethnic Global Array (v8). After genotype imputation using the TOPMed imputation server with the TOPMed r3 reference panel [28], we applied QC procedures, including imputation quality score (r^2^) > 0.7, MAF > 5%, a Hardy–Weinberg Equilibrium (HWE) P-value > 1 × 10⁻⁶, genotyping call rate > 99% per individual and per marker, and concordance between reported and genetic sex. 2,140 offspring had matched maternal CRP data, with 5,791,060 common variants available for analysis. All genotype positions were based on the GRCh38 (hg38) genome build.

### Statistical analysis

The ancestry of the 4,326 multi-ancestry maternal participants was determined using SNPweights v2.1 [29], based on reference data from the 1000 genome project consortium [30]. We assigned ancestry if the predicted ancestry proportion exceeded 80% [26], resulting in classification into six ancestral groups: African (AFR, n=349), Admixed American (AMR, n=162), East Asian (EAS, n=84), European (EUR n=2,749), South Asian (SAS, n=22), and Unassigned Ancestry (n=960). We primarily investigated the genetic contributions of first-trimester CRP in European ancestry cohort, given its larger sample size and population homogeneity. For this analysis, we restricted to unrelated individuals of European ancestry (n = 2,736) to avoid confounding due to relatedness, and performed GWAS using PLINK v2.0 [31]. For the multi-ancestry cohort, GWAS was performed using the GENESIS [32], which accounts for population structure and relatedness using a linear mixed-effects model. In both analyses, covariates included standardized age, standardized age squared, 10 principal components (PCs) for genetic ancestry, and clinical sites.

Fine-mapping was performed using SuSiE-inf [33] with in-sample linkage disequilibrium (LD) calculated using PLINK v1.9 [34]. Fine-mapping regions were defined as ±1MB around each lead variant, allowing for up to five independent signals per region. For each signal, SuSiE-inf identified a 95% credible set containing variants that together account for 95% of the posterior probability of causality. For functional downstream analysis, we used Functional Mapping and Annotation (FUMA) tool v1.6.3 [35], which requires input data aligned to the GRCh37 genome build. Therefore, the GWAS summary statistics based on GRCh38 were lifted over to GRCh37 using the UCSC liftOver tool [36] to ensure compatibility.

To estimate the genome-wide genetic correlation between CRP and GDM, we applied LDSC [37], which assesses the extent of shared genetic architecture between two traits using GWAS summary statistics. The primary analysis used summary statistics from the nuMoM2b maternal European CRP GWAS and a large GDM GWAS in Elliott et al. (2024) [38]. As a complementary analysis, we repeated LDSC using CRP summary statistics from a large population-based GWAS by Said et al. (2022) [16], paired with the same GDM dataset. Additionally, to evaluate whether genetic predisposition to elevated CRP levels contributes to GDM risk within the nuMoM2b European maternal cohort, we constructed PRS using the clumping and thresholding approach [39] in PLINK v1.9 [34]. SNP weights were derived from the same two CRP GWAS datasets described above.

Finally, to investigate the potential contribution of offspring genetic variation to maternal CRP levels, we performed an offspring-based GWAS of maternal first-trimester CRP. Due to the limited sample size of available offspring genotype data, this analysis was conducted using the full multi-ancestry sample in GENESIS, adjusting for the same covariates as in the maternal analysis, with the addition of offspring sex. The genome-wide significance (GWS) level was set at 5 × 10^-8^. The overall study and analysis workflow is illustrated in Fig 1.

**Fig 1.**
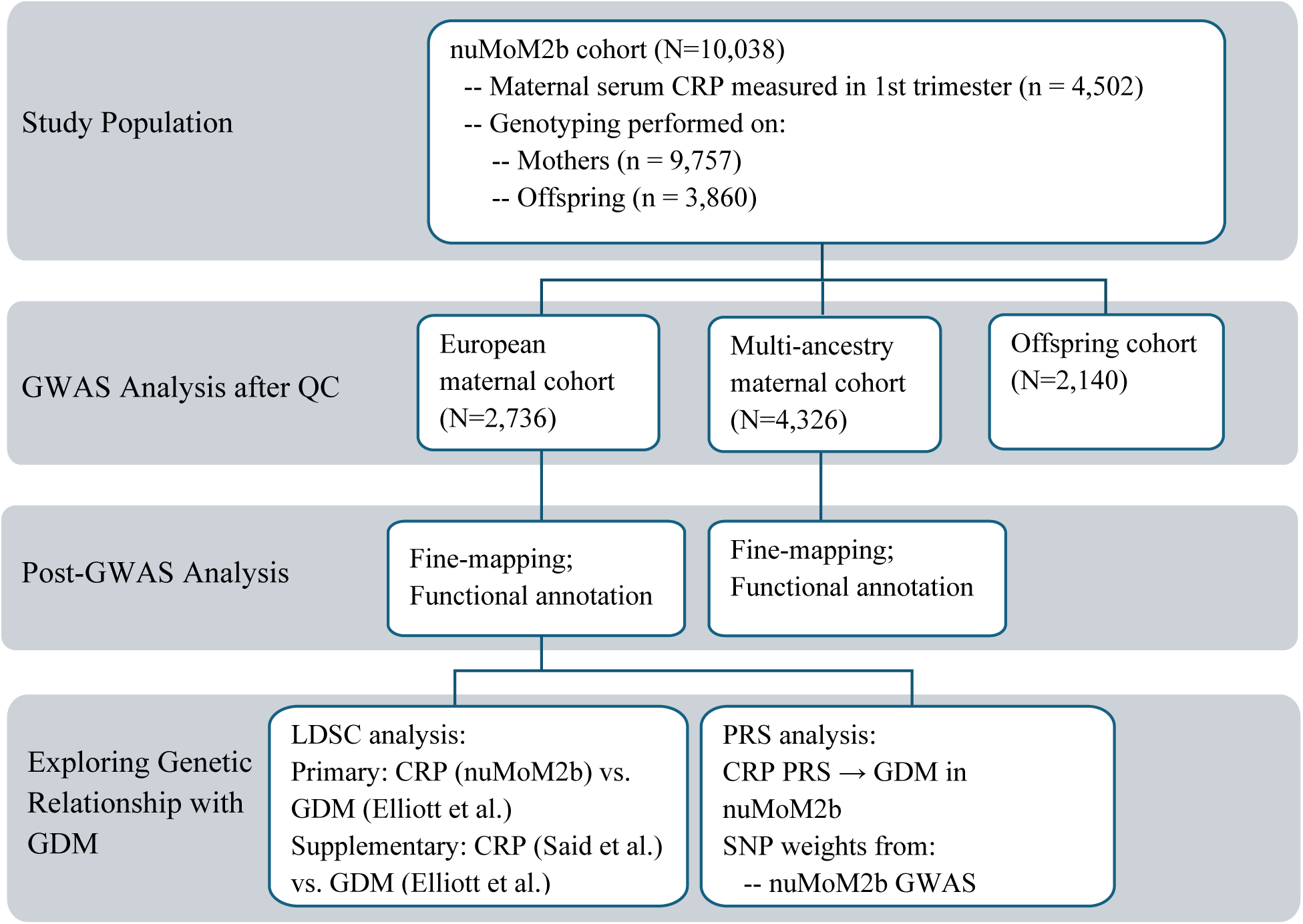
Overview of Study Design and Analysis Workflow. GDM: gestational diabetes mellitus; LDSC: LD Score Regression; PRS: polygenic risk score.

## Results from European maternal cohort

### GWAS of first-trimester CRP levels

Three GWS loci including 166 genetic variants were identified in the European maternal cohort within the nuMoM2b study (Fig 2A, S1 Table). The genomic inflation factor (λ = 1.01) suggests minimal inflation of test statistics due to population stratification or other confounding factors, as further supported by the QQ plot (S1 Fig). Additionally, principal component analysis (PCA) confirmed genetic ancestry consistency within the cohort (S2 Fig). The lead SNP among the 166 associated variants was rs1130864, an intronic variant within the *CRP* gene on chromosome 1 (β = 0.25, P = 6.94 × 10^-17^; Fig 3B). A second locus on Chromosome 1 was marked by lead SNP rs12130672, located near *LEPR* (β = −0.16, P = 1.31 × 10^-8^; Fig 3A). The third locus, on chromosome 12, spans the *HNF1A* and *C12orf43* genes and is marked by lead SNP rs9738226, which lies within *HNF1A* (β = −0.20, P = 1.24 × 10^-12^; Fig 3C). Fine-mapping analysis identified four 95% credible sets (S2 Table). Three of these overlapped with the genome-wide significant loci identified in the GWAS. One credible set, however, included SNPs that did not reach genome-wide significance but were prioritized based on high posterior probabilities. These SNPs are located within *ENSG00000297913*, a long non-coding RNA (lncRNA) positioned antisense to the *CRP* gene.

**Fig 2.**
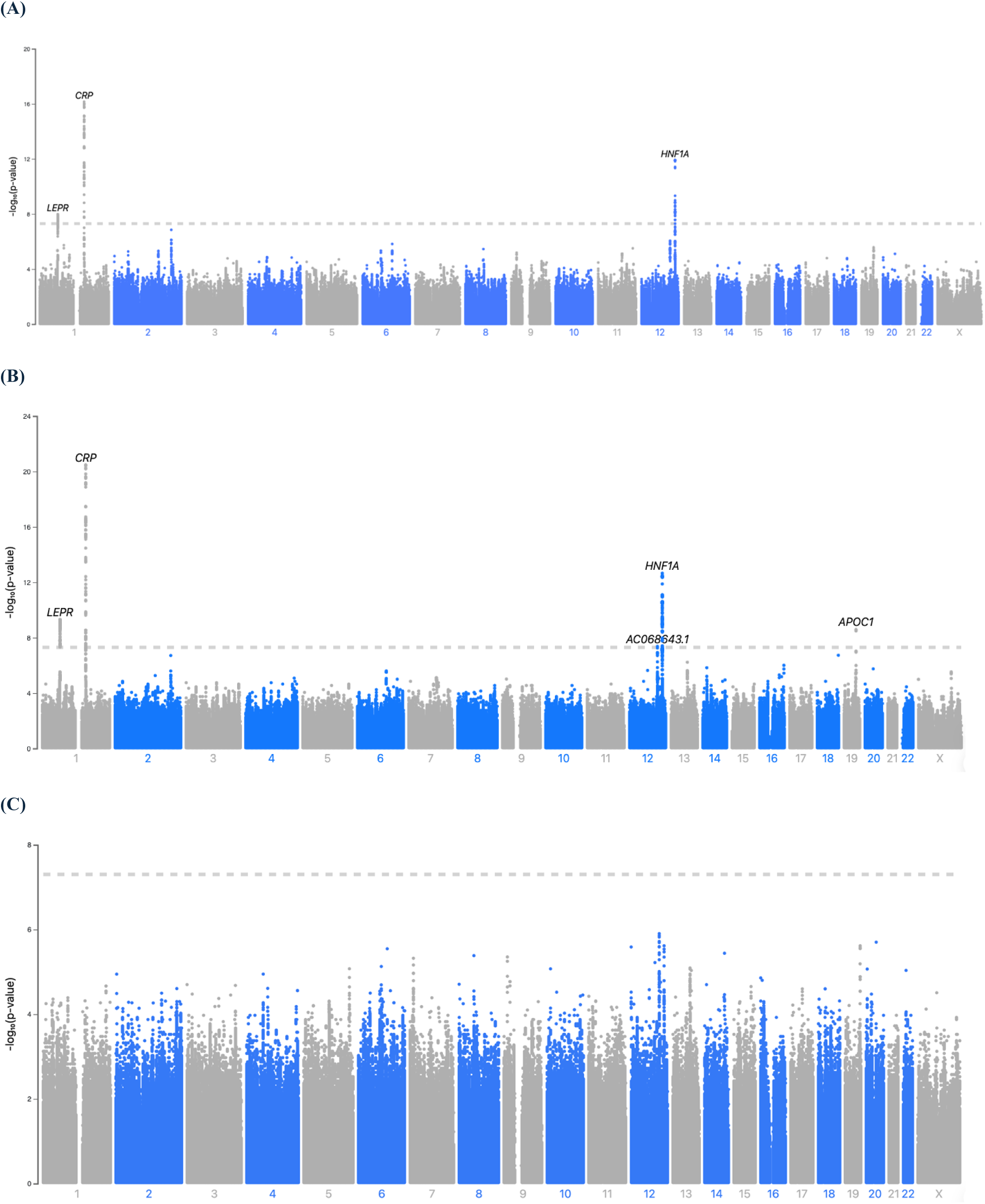
Manhattan plot of GWAS of first-trimester CRP. -log10 P values on the y axis are plotted against physical position (GRCh38) on the x axis. The horizontal dashed line represents the significance level (P < 5 × 10^-8^). (A) European maternal cohort. (B) multi-ancestry maternal cohort. (C) multi-ancestry offspring cohort.

**Fig 3.**
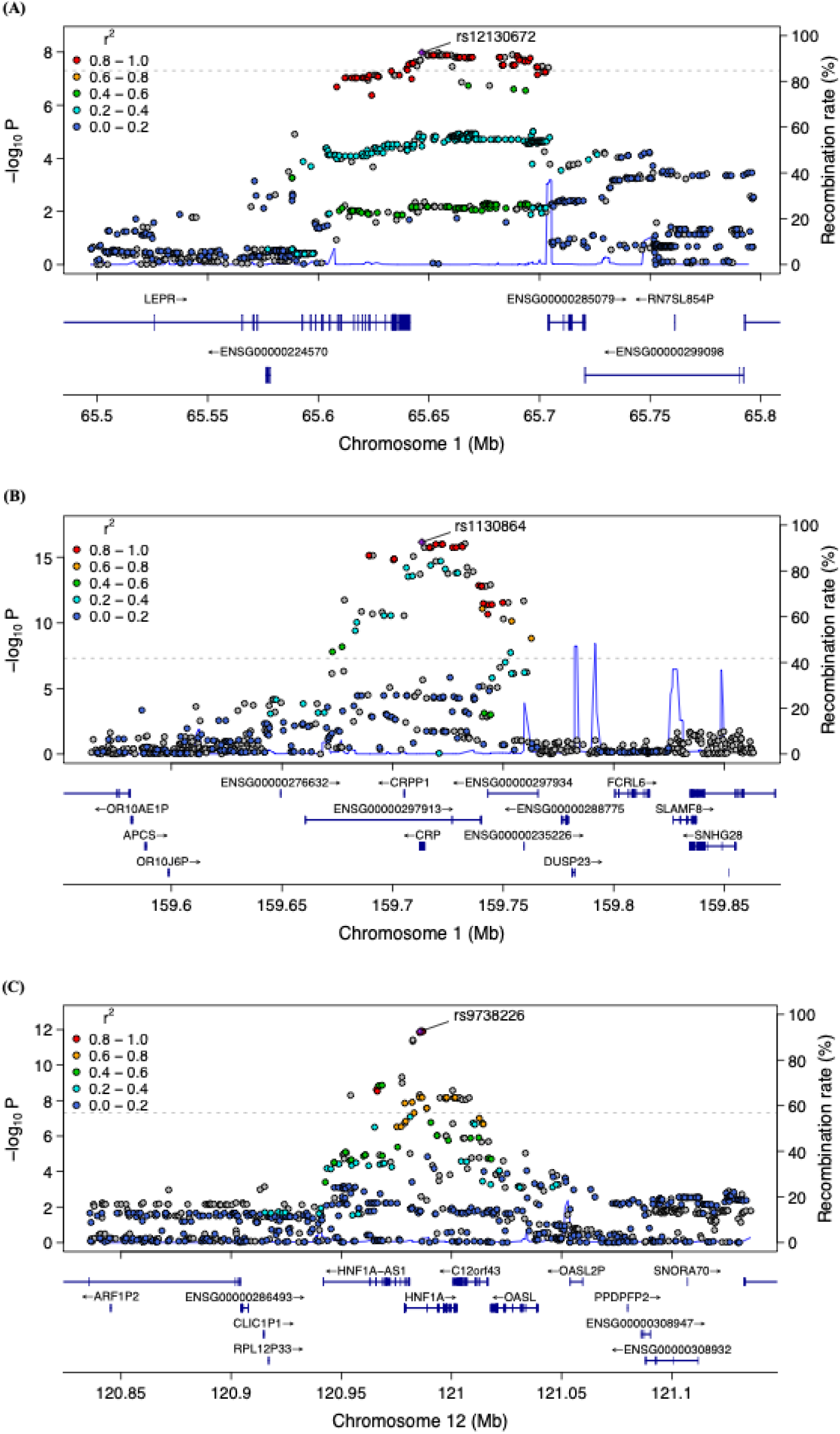
Regional plots of genome-wide significant loci in the European maternal cohort. (A) Regional plot for SNP rs12130672. (B) Regional plot for SNP rs1130864. (C) Regional plot for SNP rs9738226. The lower panel of each plot shows the relative location of nearby genes and their transcriptional direction. The lead SNP is highlighted as a purple diamond, while surrounding SNPs are color-coded based on their linkage disequilibrium (LD, r²) with the lead SNP, calculated using the 1000 Genomes European reference pane.

### Functional downstream analysis

Functional annotation of GWS variants and their LD-linked variants (S3 Table) revealed two variants on chromosome 1: rs876537 (ncRNA_exonic *CRPP1*) and rs12083620 (intergenic), both with combined annotation-dependent depletion (CADD) scores above 12.37. Regulatory annotation using RegulomeDB score identified rs4656849 (intergenic), rs7953249 (intergenic) and rs1169310 (UTR3 *HNF1A*) as variants that may affect transcription factor (TF) binding or gene expression. Furthermore, FUMA ANNOVAR analysis showed that over 80% of GWS SNPs and their LD-linked variants were in intergenic and intronic regions (S3 Fig). Those GWS variants were also mapped to 98 protein-coding genes (S4 Table) via the positional mapping, expression quantitative trait loci (eQTL), and chromatin interaction mapping. Not only the proximal genes but also the distal genes were identified with eQTL/chromatin interaction mapping (S4 and S5 Figs). All the variants in GWAS were annotated to 19,273 protein coding genes using FUMA-MAGMA, of which three genes (*CRP*, *HNF1A*, *C12orf43*) are associated with first-trimester CRP at Bonferroni significance level (P < 2.60 × 10^-6^) (S6 Fig, S5 Table). FUMA-MAGMA tissue expression analysis identified enrichment of first-trimester CRP associated genes in kidney, fallopian tube, pancreas, and heart tissues (S7 Fig), although they did not reach Bonferroni-corrected statistical significance (P ≤ 1.67 × 10^-3^).

### Genetic relationship between CRP and GDM

SNP-based heritability of first-trimester CRP was estimated at 30.3% (SE = 16.4%). We first assessed genome-wide genetic correlation between CRP and GDM using LDSC applied to our first-trimester CRP GWAS in nuMoM2b alongside the external GDM GWAS in Elliott et al. (2024). This pregnancy-specific analysis revealed no significant genetic correlation (r_g_ = 0.044, SE = 0.221, P = 0.841), indicating minimal shared heritability when CRP is measured in early gestation. As a complementary analysis, we calculated the genetic correlation between CRP measured in the general population (Said et al. (2022)) and GDM using LDSC. We observed a significant genetic correlation (r_g_ = 0.197, SE = 0.058, P = 0.0007), suggesting a moderate shared genetic architecture between GDM and systemic inflammation in the general population. Finally, to test whether the genetic predisposition to CRP influences GDM risk within nuMoM2b, we constructed polygenic risk scores (PRS) with SNP weights from both CRP GWAS sources, respectively, and modeled their association with GDM status in the nuMoM2b cohort. Neither PRS was significantly associated with GDM (P > 0.05).

## Results from multi-ancestry maternal cohorts

### GWAS of first-trimester CRP levels

In the multi-ancestry maternal cohort, five GWS loci including 237 genetic variants were identified in multi-ancestry cohort (Fig 2B, S6 Table). The genomic inflation factor (λ = 1.01) and corresponding QQ plot (S8 Fig) suggest minimal inflation of association statistics, indicating limited population stratification or other confounding influences. The strongest association was observed for rs1341665, an intergenic variant on chromosome 1q23.2 near the *CRP* gene (β = - 0.23, P = 3.36 × 10^-21^; Fig 4B). On the same chromosome, the top variant rs4311928 near *LEPR* also reached genome-wide significance (β = −0.14, P = 5.12 × 10^-10^; Fig 4A). Two additional loci were identified on chromosome 12, marked by lead variant rs7298698 within *ENSG00000257703* (also known as *AC068643.1*) (β = 0.12, P = 4.27 × 10^-8^; Fig 4C) and rs7979473, an intronic variant in *HNF1A* (β = −0.17, P = 2.21 × 10^-13^; Fig 4D). The final significant locus was located on chromosome 19, marked by rs12721051, an intronic variant in *APOC1* (β = −0.19, P = 2.59 × 10^-9^; Fig 4E). Fine-mapping analysis identified six 95% credible sets across the five GWS loci (S7 Table). One of the credible sets consisted of a single variant (rs2369146) that did not overlap with a GWS locus but was highly prioritized based on posterior probability.

**Fig 4.**
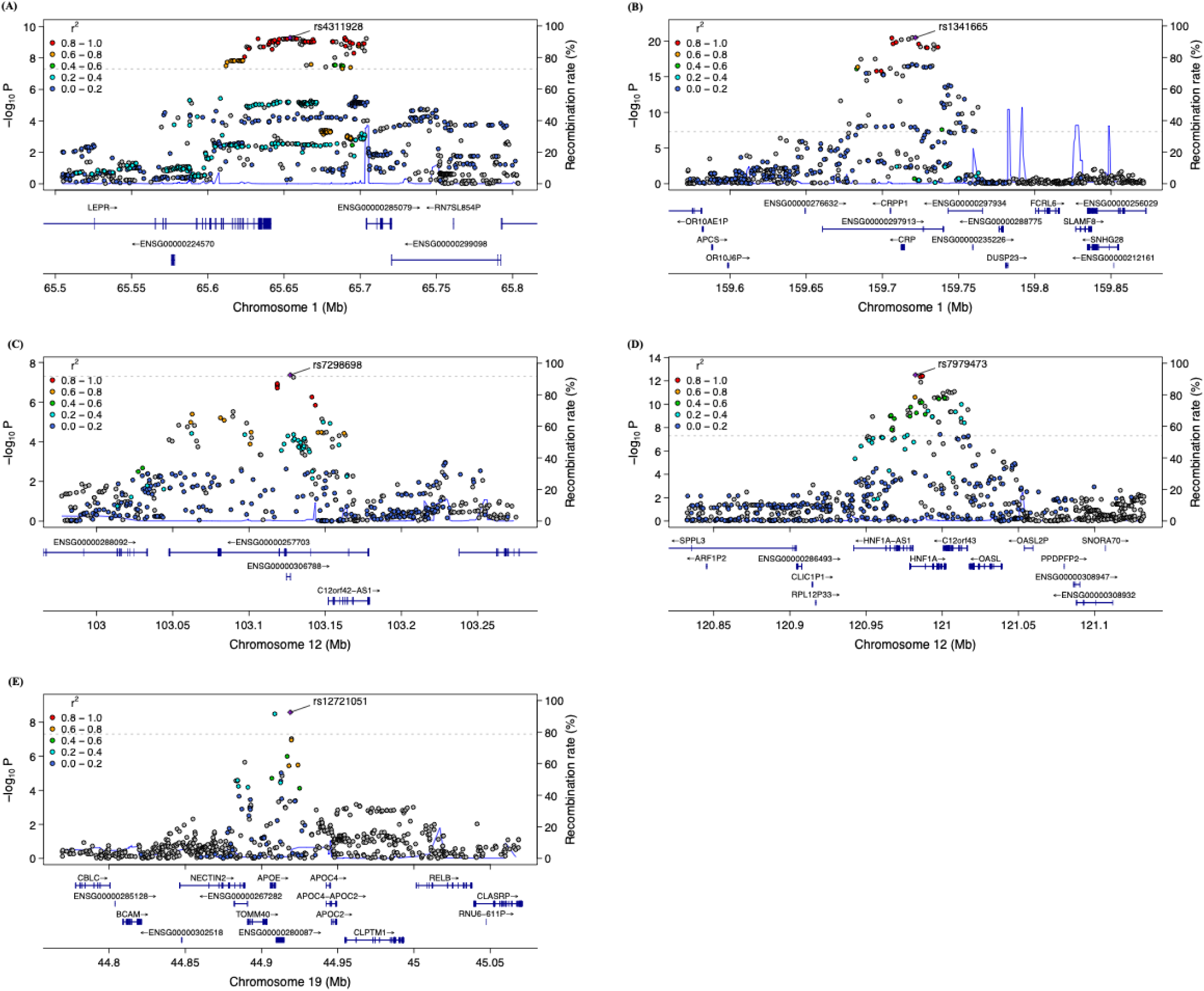
Regional plot for genome-wide significant loci in the multi-ancestry maternal cohort. (A) Regional plot for SNP rs4311928. (B) Regional plot for SNP rs1341665. (C) Regional plot for SNP rs7298698. (D) Regional plot for SNP rs7979473. (E) Regional plot for SNP rs12721051. The lower panel of each plot shows the relative location of nearby genes and their transcriptional direction. The lead SNP is highlighted as a purple diamond, while surrounding SNPs are color-coded based on their linkage disequilibrium (LD, r²) with the lead SNP, calculated using the 1000 Genomes reference panel (ALL populations).

### Functional downstream analysis

Functional annotation of GWS variants and their LD-linked variants (S8 Table) identified 11 prioritized variants with CADD scores above 12.37. Regulatory annotation using RegulomeDB scores indicated six variants with potential regulatory roles, including four intergenic variants (rs4656849, rs6489786, rs7953249, rs2251468), one variant in the 3’ UTR of *HNF1A* (rs1169310) and one intronic variant in *C12orf43* (rs1169314), all of which may affect TF binding and chromatin activity. FUMA ANNOVAR results showed that over 80% of GWS SNPs and their LD-linked variants were in intergenic and intronic regions (S9 Fig). Those GWS variants were mapped to 119 protein-coding genes (S9 Table) via the positional mapping, expression quantitative trait loci (eQTL) and chromatin interaction mapping. Not only the nearby genes but also the distal genes were identified with eQTL/chromatin interaction mapping (S10-S12 Figs). All the variants in GWAS were annotated to 19366 protein coding genes with FUMA-MAGMA, of which five genes (*CRP*, *HNF1A*, *C12orf43*, *APOE*, *APOC1*) are associated with first-trimester CRP at Bonferroni significance (P < 2.58×10^-6^) (S13 Figs, S10 Table). FUMA-MAGMA tissue expression analysis identified enrichment of first-trimester CRP associated genes in liver tissues (S14 Fig), although they did not reach statistical significance at the Bonferroni cut-off (P < 1.67×10^-3^).

## Results from offspring cohorts

In the offspring GWAS of maternal CRP levels, no variants reached genome-wide significance (P < 5×10^-8^, Fig 2C). However, 42 variants demonstrated suggestive associations at a less stringent threshold (P < 1×10^-5^, S11 Table). Notably, some of these variants were located in or near genes previously implicated in inflammatory processes, such as *HNF1A* and *FBLN5* [40]. An interactive plot showing all association results is available at https://my.locuszoom.org/gwas/130031/?token=4c1783ffb536418da22d63abaeb7d305 for further exploration.

## Discussion

In this comprehensive GWAS investigating the genetic contributions of CRP levels during early pregnancy, we analyzed both maternal and offspring genotypes from the nuMoM2b cohort. Consistent with previous large-scale CRP GWAS conducted in general European populations [16], we identified three GWS loci in the European maternal cohort, including variants within or near *CRP*, *LEPR*, *HNF1A* and *C12orf43* genes. These loci are well-established regulators of systemic inflammation and metabolic homeostasis [41,42]. For example, LEPR, the leptin receptor, is ubiquitously expressed receptor involved in regulating energy balance, appetite, and immune function [43,44]. Polymorphism or mutation in LEPR gene are associated with changes in diet and obesity [45,46]. HNF1A, the (hepatocyte nuclear factor 1 homeobox A), is a transcription factor that is expressed in many tissues including pancreas, liver, and kidney [47]. Mutation in HNF1A is also known to cause a monogenic form of diabetes and polymorphism has been linked to risk of GDM [48–51]. While the number of genome-wide significant variants was much smaller than those reported in the large general European study, the overall consistency in locus-level associations reinforces the robustness of our findings. Fine-mapping analyses further identified additional candidate variants, including those in non-coding regions such as ENSG00000297913, a lncRNA transcribed antisense to the CRP gene. Antisense lncRNAs can regulate gene expression through transcriptional interference, RNA stability, or chromatin remodeling mechanisms [52], suggesting that these variants may modulate CRP levels even in the absence of genome-wide significance.

Notably, two additional GWS loci—marked by lead SNPs rs7298698 (within *ENSG00000257703*) and rs12721051 (within APOC1)—were identified in the multi-ancestry cohort beyond those detected in the European analysis. Although both variants had similar effect sizes and allele frequencies across the two cohorts (e.g., rs7298698: β = 0.124, MAF = 0.48 in multi-ancestry vs. β = 0.137, MAF = 0.64 in European; rs12721051: β = −0.189, MAF = 0.144 in multi-ancestry vs. β = −0.171, MAF = 0.173 in European), they reached genome-wide significance only in the larger, multi-ancestry sample, likely due to increased statistical power. This highlights the importance of diverse cohorts in uncovering associations that may be missed in ancestry-restricted analyses. Functionally, this region on chromosome 19 includes both *APOC1* and *APOE*, which are closely linked and have been implicated in lipid metabolism, inflammation, and cardiovascular risk [53]. In contrast, ENSG00000257703 encodes a long non-coding RNA with unclear biological function but consistent association signals in CRP GWAS [16], suggesting a possible regulatory role that needs further investigation.

Although no loci reached genome-wide significance in the offspring GWAS of maternal CRP during pregnancy, several loci exhibited suggestive associations (P < 1×10^-5^), raising the possibility of offspring genetic contributions to maternal inflammatory status during pregnancy.

This concept is supported by prior studies demonstrating offspring genetic effects on maternal phenotypes, including blood pressure and glucose metabolism [20–22]. Such effects are thought to arise from placental signaling or fetal–maternal immunologic interactions during gestation. Although our findings were not statistically significant, they may reflect early evidence of such offspring genetic influence on maternal CRP. However, the limited power due to the relatively small offspring samples likely constrained our ability to detect significant associations. Future studies with larger offspring genotype data will be crucial to further elucidate these contributions.

Finally, genetic correlation analysis using our first-trimester CRP GWAS in nuMoM2b revealed no evidence of shared heritability with GDM. In contrast, a complementary analysis using a much larger, population-based CRP GWAS found a modest correlation. Clinically, the modest genetic correlation observed in the general population suggests that CRP may contribute to GDM risk through shared genetic pathways. This discrepancy may reflect differences in CRP’s genetic architecture during early pregnancy versus in the general population. Alternatively, it could be due to a smaller sample size in nuMoM2b study that lacks the power to detect shared variants driving GDM risk. Additionally, our analysis was restricted to common variants (MAF > 0.05) and therefore potentially not capturing rare variants. Furthermore, the PRS of CRP was not statistically associated with GDM in nuMoM2b. These findings suggest that although early pregnancy CRP levels are phenotypically linked to GDM [11], the genetic predisposition to higher CRP does not translate into increased GDM risk. This supports the interpretation of CRP as a downstream biomarker of inflammatory or metabolic changes that accompany GDM, rather than a causal driver of disease. The lack of genetic correlation and PRS association implies that shared genetic etiology between CRP and GDM is limited, and that the observed associations may instead reflect physiological adaptations of pregnancy, subclinical inflammation, insulin resistance, or other environmental exposures such as obesity, diet, or stress. Thus, CRP may serve as a useful clinical indicator of underlying risk, but not a mechanistic target for GDM prevention.

While the modest sample size of nuMoM2b limits power to detect variants with smaller or rare effects on first-trimester CRP levels, the study’s strength lies in its uniquely rich design. nuMoM2b is one of the largest U.S. prospective pregnancy cohorts with biospecimens collected in the first trimester and includes both maternal and offspring genotypes across multiple ancestries. This enables investigation of pregnancy-specific genetic architecture of CRP—a rare opportunity not captured in large general population studies. These features make the dataset uniquely valuable for advancing our understanding of maternal inflammation during early pregnancy.

In conclusion, by validating known systemic - inflammation loci (e.g., *CRP*, *LEPR*, *HNF1A*, *APOC1*) in pregnant women, exploring suggestive signals in offspring, we deliver the first gestation-tailored genetic map of CRP regulation. These results not only fill a critical gap in the literature but also supply pregnancy-specific data for future meta-analyses and PRS construction, laying essential groundwork for subsequent studies with larger and more diverse cohorts.

## Data Availability Statement

Access to the nuMoM2b individual-level data is available through dbGaP under study accession phs002808.v1.p1 and the NICHD repository DASH.

Interactive visualizations of the GWAS results can be accessed at: European maternal GWAS https://my.locuszoom.org/gwas/612155/?token=cd3ac564de804ec9afb1ee733e345f4c;

Multi-ancestry maternal GWAS https://my.locuszoom.org/gwas/833469/?token=017c786d7c8640c8913c5c87b75378c9;

Offspring GWAS https://my.locuszoom.org/gwas/130031/?token=4c1783ffb536418da22d63abaeb7d305.

## Acknowledgements

This research was supported in part by Lilly Endowment, Inc., through its support for the Indiana University Pervasive Technology Institute.

DNA extraction and genotyping funded by the Indiana University Grand Challenge Precision Health Initiative.

This study is supported by grant funding from the Eunice Kennedy Shriver National Institute of Child Health and Human Development (NICHD): U10 HD063036, K08 HD109636, RTI International; U10 HD063072, Case Western Reserve University; U10 HD063047, Columbia University; U10 HD063037, Indiana University; U10 HD063041, University of Pittsburgh; U10 HD063020, Northwestern University; U10 HD063046, University of California Irvine; U10 HD063048, University of Pennsylvania; and U10 HD063053, University of Utah. In addition, support was provided by respective Clinical and Translational Science Institutes to Indiana University (UL1TR001108) and University of California Irvine (UL1TR000153).

## Author contributions

Yu Zhang: Conceptualization, Data curation, Formal analysis, Methodology, Project administration, Software, Visualization, Writing – original draft, Writing – review & editing; Amy Moore: Conceptualization, Writing – review & editing; Kelli K. Ryckman: Conceptualization, Writing – review & editing; Qi Yan: Resources; Writing – review & editing; Rafael F. Guerrero: Resources; Writing – review & editing; Ming Li: Writing – review & editing; David M. Haas: Resources; Writing – review & editing; Robert M. Silver: Writing – review & editing; Lynn M Yee: Writing – review & editing; Uma M. Reddy: Writing – review & editing; Judith Chung: Writing – review & editing; Kok Lim Kua: Conceptualization, Supervision, Writing – review & editing; Nianjun Liu: Funding Acquisition, Supervision, Writing – review & editing

